# Hypertension among people living with HIV/AIDS in Cameroon: A cross-sectional analysis from Central Africa International Epidemiology Databases to Evaluate AIDS

**DOI:** 10.1101/2020.08.16.20176008

**Authors:** Anastase Dzudie, Don Hoover, Hae-Young Kim, Rogers Ajeh, Adebola Adedimeji, Qiuhu Shi, Walter Pefura Yone, Denis Nsame Nforniwe, Kinge Thompson Njie, Andre Pascal Kengne, Vanes Peter Ebasone, Blaise Barche, Zoung-Kany Bisseck Anne Cecile, Denis Nash, Marcel Yotebieng, Kathryn Anastos

**Author notes:** Corresponding author: Professor Anastase Dzudie, *Service of Internal Medicine and Cardiology, Douala General Hospital, Cameroon* Po box 4856, Douala, Cameroon.

## Abstract

**Background:** Antiretroviral therapy (ART) success has led people to live longer with HIV/AIDS (PLWH) and thus be exposed to increasing risk of cardiovascular diseases (CVD). Hypertension (HTN), the biggest contributor to CVD burden, is a growing concern among PLWH. The current report describes the prevalence and predictors of HTN among PLWH in care in Cameroon.

**Methods:** This cross-sectional study included all PLWH aged 20 years and above who received care between 2016 and 2019 at one of the three Central Africa International Epidemiology Databases to Evaluate AIDS (CA-IeDEA) sites in Cameroon (Bamenda, Limbe, and Yaoundé). HTN was defined as blood pressure (BP) ≥140/90 mm Hg or self-reported use of antihypertensive medication. Logistic regressions models examined the relationship between HTN and clinical characteristics, and HIV-related factors.

**Results:** Among 9,839 eligible PLWH, 66.2% were female and 25.0% had prevalent HTN [age-standardized prevalence 23.9% (95% CI: 22.2–25.6)], among whom 28 (1.1%) were on BP lowering treatment, and 6 of those (21.4%) were at target BP levels. Median age (47.4 vs. 40.5 years), self-reported duration of HIV infection (5.1 vs 2.8 years), duration of ART exposure (4.7 vs 2.3 years), and CD4 count (408 vs 359 cell/mm^3^) were higher in hypertensives than non-hypertensives (all p< 0.001). Age and body mass index (BMI) were independently associated with higher prevalent HTN risk. PLWH starting ART had a 30% lower risk of prevalent HTN, but this advantage disappeared after a cumulative 2-year exposure to ART. There was no significant association between other HIV predictive characteristics and HTN.

**Conclusion:** About a quarter of these Cameroonian PLWH had HTN, driven among others by age and adiposity. Appropriate integration of HIV and NCDs services is needed to improve early detection, treatment and control of common comorbid NCD risk factors like hypertension and safeguard cardiovascular health in PLWH.

## INTRODUCTION

The advent and widespread use of antiretroviral therapy (ART) has successfully achieved viral suppression in 76% of sub-Saharan Africans living with HIV/AIDS [1]. Since 2005, there has also been a decline in AIDS-related deaths by 35% globally and by 39% in sub-Saharan Africa (SSA). This decline is partly attributed to new recommendations which entails providing antiretroviral therapy (ART) for all people living with HIV (PLWH) at diagnosis irrespective of CD4 counts, otherwise known as “*Treat All*”[2]. These improvements have narrowed the gap in life expectancy between persons with and without HIV [3][4]. However, increases in life expectancy among PLWH are accompanied by increased occurrence of non-communicable diseases (NCDs), particularly cardiovascular disease (CVD) [5][6].

Hypertension (HTN) remains the leading risk factor for CVD which is the main cause of death globally, accounting for 10.4 million deaths annually [7][8]. In 2015, the World Health Organization (WHO) indicated that the number of people with HTN in the general population was highest in sub-Sahara Africa (SSA), at about 46% of adults aged 25 years and older, compared to 35 to 40% elsewhere [8]. In PLWH, previous studies in Africa and elsewhere showed a high prevalence of HTN [6,9] with some studies estimating that one-quarter of PLWH have HTN [10].

In SSA, reported HTN prevalence in PLWH ranged from 12.5% to 28.5%, including in Cameroon where two single center cross-sectional studies of small sample size estimated that 25% and 28% of PLWH were hypertensive in 2015 and 2016 [11–13]. In a systematic narrative review, Nguyen et al estimated that HTN prevalence in PLWH from low- and middle-income countries ranged from 8.7 to 45.9%[14], suggesting that HTN burden in PLWH populations has not been well-established or otherwise may vary greatly. Accurate estimates of HTN burden in SSA PLWH is needed to guide policies that will facilitate optimization of both HIV and HTN in HIV clinics as recommended by the Pan African Society of cardiology [15].

Cameroon bears the largest burden of HIV/AIDS in Central Africa and one of the highest in SSA with an estimated number of PLWH around 540,000 in 2018 [16]. According to the Cameroon Population-Based HIV Impact Assessment study, 52% of this HIV-infected population was receiving ART and had access to healthcare services in 2017[17]. However, HIV care is generally provided in dedicated HIV clinics while patients with NCDs receive care in general medical services, with little or no overlap in management of HIV and NCDs, particularly HTN, limiting the assessment of the true burden of NCDs in PLWH. Similarly, because of the lack of integration of NCDs and HIV care services, data are limited on access to HTN treatment and outcomes among PLWH in SSA in general and in Cameroon in particular. Such information would help in allocation of resources, and development of cost-effective therapeutic strategies and programs for integrating HIV and HTN management. We therefore sought to examine the prevalence and predictors of HTN among PLWH attending three large clinics in Cameroon.

## METHODS

### Study design and setting

This cross-sectional study used data from the International Epidemiology Databases to Evaluate AIDS (IeDEA) in Cameroon. Cameroon, Burundi, the Democratic Republic of Congo, the Republic of Congo and Rwanda, comprise central Africa-IeDEA (CA-IeDEA) contributing secondary data from medical records to the regional data center, one of the seven that compose the global IeDEA consortium [18]. Each regional data center collaborates with clinical sites to identify and define key variables, harmonize and effectively analyze the data, and generate large datasets. In Cameroon, three clinical sites participate in the CA-IeDEA: the Limbe Regional Hospital (LRH), the Bamenda Regional Hospital (BRH), and the Yaoundé Jamot Hospital (YJH). At these public health sites, ART and care delivery is supported by the Cameroon National AIDS Control Committee.

### Population, enrolment into IeDEA

In Cameroon, the National Ethical Committee requires a written consent from participants before their data are extracted from medical records into the IeDEA database. Starting in January 2016, all PLWH in care at the HIV clinic in each of the three sites were approached during a routine clinic visit and invited to participate in IeDEA and consented. During the consenting visit (enrolment into IeDEA), additional demographic and clinical variables were collected to supplement routine data, including blood pressure (BP), weight and height.

For this study, we included all non-pregnant (at enrolment) HIV-positive participants aged 20 years or older enrolled into IeDEA from January 2016 through December 2019. We excluded patients with missing systolic and diastolic BP measurements within the first six months of enrolment into IeDEA.

Patient data are routinely collected by research assistants on case report forms. Data are then captured from paper forms into REDCap[19,20], a free, secure and flexible web-based clinical research data capture platform. These data included demographic characteristics, clinical conditions including weight, height, and systolic and diastolic blood pressure (BP), HIV diagnosis, duration and disease control, laboratory test results, medication use (antiretroviral (ARV) and non-ARV). During the study period, the standard first-line regimen for PLWH in Cameroon was a combination of three drugs including two nucleoside or nucleotide reverse transcriptase inhibitors (NRTI) like zidovudine (AZT) or lamivudine (3TC) or Tenofovir (TDF) and a nonnucleoside reverse transcriptase inhibitor (NNRTI) like efavirenz (EFV) or nevirapine (NVP) as either AZT/3TC/EFV, AZT/3TC/NVP, TDF/3TC/EFV or TDF/3TC/NVP. Second-line regimens during this period included protease inhibitor-(PI) and Integrase Strand Transfer Inhibitor (INSTI) based therapy.

### Blood pressure measurement

Blood pressure measurements were obtained during the enrolment visit by trained healthcare professionals, using validated automated BP monitors and following a standardized protocol for office BP measurement. Briefly, patients were seated comfortably in a quiet office for 5 min. BP measurements were done on the right arm using an OMRON M3 HEM-7131-E device (Omron Healthcare Co Ltd, Kyoto, Japan) and appropriate cuff size. Three BP measurements were recorded 1–2 min apart, and an average of the two last readings used in the analysis.

### Definition of outcomes

Our primary outcome was HTN at enrolment in IeDEA. HTN was defined following the European Society of Cardiology(ESC)/European Society of HTN (ESH) guidelines for HTN[21] as Systolic BP (SBP) ≥140mmHg and/or diastolic BP (DBP) ≥90mmHg and/or current use of antihypertensive medication from medical records. HTN was further graded following the ESC/ESH guidelines of HTN as optimal for SBP < 120 mmHg and DBP < 80 mmHg, normal for SBP 120–129 mmHg and/or DBP 80–84 mmHg, high normal for SBP 130–139 mmHg and/or DBP 85–89 mmHg, grade 1 HTN for SBP 140–159 mmHg and/or DBP 90–99 mmHg, grade 2 for SBP 160–179 mmHg and/or DBP 100–109 mmHg, grade 3 for SBP ≥180 mmHg and/or DBP ≥110 mmHg and isolated systemic hypertension for SBP ≥140 mmHg and DBP < 90 mmHg [21]. Secondary outcomes included the proportion of all hypertensive participants receiving treatment for HTN at the time of enrolment into IeDEA and controlled HTN defined as BP < 140/90mmHg in those on antihypertensive treatment.

### Potential predictors of hypertension

Values of all predictors investigated were collected at time of enrolment. Candidate predictors included age (categorized into 18–29, 30–49, ≥50 years), sex (male vs female), marital status (single, married, live with a partner, separated or divorced and widowed), education level (none, primary, secondary up to high school, high school or more), employment status (employed and unemployed), smoking status (never, current, former) and alcohol consumption (never, monthly or less, 2-3 standard drinks per months, ≥ one per standard drink week). A standard alcoholic drink was considered as equivalent to 1 can (340mL) of beer, 1 glass (125mL) wine, or 1 shot (25mL) of spirits.

The following clinical parameters were also considered as candidate predictors: body mass index (BMI, kg/m^2^, categorized as underweight ≤18, normal 18–24.9, mildly overweight 25–29.9, overweight 30–34.9, or obese ≥35 kg/m^2^); blood glucose categorized based on fasting plasma glucose (FPG) as normal (FPG< 100 mg/dL), impaired fasting glucose (FPG between 100 and 125 mg/dL) and diabetes (FPG ≥126 mg/d or ongoing antidiabetic medications). HIV disease categories were based on the WHO classification [22]. ART exposure was categorized as having received ART prior to the date of enrollment in study. Estimated duration on ART among users was the time from the first prescription of ART to the date of enrolment BP measurement. Diagnosed duration of HIV infection was estimated time from first recorded or self-reported HIV positive test to the date of enrollment into CA-IeDEA project.

### Statistical analysis

The crude prevalence of HTN was calculated among study participants with available BP measurement, while the proportion receiving treatment was calculated among all hypertensive participants and the proportion with controlled calculated among those receiving treatment. Age-standardized sex-specific prevalence was calculated using the age distribution of PLWH in the 2018 distribution of the Cameroon HIV population [23]. Medians and quartiles (Q1 = 25th percentile, Q3 = 75th percentile) and proportions were used to summarize continuous and categorical variables, respectively. Chi-square (χ^2^) tests and equivalents compared proportions, Student t-test, Analysis of the variance (ANOVA) or rank tests as appropriate compared continuous variables. Logistic regression models estimated the odds ratios (OR) and 95% confidence intervals (95% CI) as measure of the association between candidate predictors and HTN. Only variables found to be associated with the outcome in crude analysis (p< 0.05) were included in the multivariable logistic models, but duration of HIV infection was excluded due to collinearity with age and duration of ART use. The limited number of hypertensive participants on antihypertensive treatment precluded similar investigations for the two secondary outcomes. Data were analysed using SAS Version 9.4 (Cary, NC). A p-value<0.05 was considered statistically significant.

## Results

### Cohort description

Of 9,988 PLWH eligible for study inclusion, 149 (1.5%) were excluded due to missing data on BP measurement (Figure 1). The remaining 9,839 participants were included in this analysis. Table 1 depicts the distribution of participants’ characteristics overall and by HTN status. In all, 6,502 (66.2%) were women, the median age was 42 years (Q1, Q3: 35–50), 3,225 (32.9%) were single; only 1,720 (17.5%) had high school training or more and 6,245 (64.8%) were unemployed. Median time from HIV diagnosis to enrollment into IeDEA was 3.4 years (Q1, Q3: 0.2–8.1), 6,645 (67.5%) participants were on ART at enrollment with 6,312 (95.4%) on NNRTI-, 405 (6.1%) on PI- and only 4 (0.1%) on INSTI-based regimen. Among those on ART, median duration of use was 2.9 years (Q1, Q3: 0.0–7.6). Poor general health status described by WHO clinical stage III and IV was noted in 1,249 (29.9%) and 419 (10.0%) respectively. Median CD4 count was 373 cells/mm^3^ (Q1, Q3: 184–573), median systolic and diastolic BP were 122 (Q1, Q3: 107–133) and 76 (Q1, Q3: 69–86) mmHg, respectively, median BMI was 24.1 (Q1, Q3: 21.4–27.7) kg/m^2^ and median hemoglobin 11.2 (Q1, Q3: 9.8–12.6) g/dl.

**Table 1.** Demographic and clinical characteristics of people leaving with HIV/AIDS at enrolment with and without hypertension.

| Characteristics | Total<br>n=9839 | Hypertension<br>n=2450 | No<br>hypertension<br>n=7389 | p-value |
| --- | --- | --- | --- | --- |
| **Age (years)** |  |  |  |  |
| Median (25th-75<sup>th</sup> percentiles) | 42.0 (35.0,50.0) | 47.4<br>(40.1,55.0) | 40.5 (34.0-48.0) | <0.001 |
| (Min, Max) | 18.1 -105.8 | 19.0-87.1 | 18.1-105.8 |  |
| **Age category, n (%)** |  |  |  |  |
| 18-29 | 1146 (11.16) | 106 (4.3) | 1040 (14.1) | <0.001 |
| 30-39 | 3053 (31.0) | 505 (20.6) | 2548 (34.5) |  |
| 40-49 | 3220 (32.7) | 860 (35.1) | 2360 (31.9) |  |
| ≥50 | 2420 (24.6) | 979 (40.0) | 1441(19.5) |  |
| **Gender (Female), n (%)** | 6502 (66.2) | 1555 (63.5) | 4947 (67.1) | 0.001 |
| **Marital Status, n (%)** |  |  |  |  |
| Single | 3225 (32.9) | 615 (25.2) | 2610 (35.4) | <0.001 |
| Married | 3287 (33.5) | 916 (37.5) | 2371(32.2) |  |
| Live with a partner | 872 (8.9) | 148 (6.1) | 724 (9.8) |  |
| Separated/Divorced | 695 (7.1) | 193 (7.9) | 502 (6.8) |  |
| Widowed | 1734(17.7) | 572(23.4) | 1162(15.8) |  |
| **Level of education, n (%)** |  |  |  |  |
| Never went to school | 959 (9.8) | 271(11.1) | 688 (9.3) | 0.0021 |
| Primary | 4256 (43.4) | 1059 (43.4) | 3197(43.4) |  |
| Secondary | 2875 (29.3) | 655 (26.9) | 2220 (30.1) |  |
| High school or more | 1720 (17.5) | 454 (18.6) | 1266 (17.2) |  |
| **Employment status, n (%)** |  |  |  |  |
| Employed | 3542 (35.2) | 856 (35.2) | 2686 (35.5) | 0.228 |
| Unemployed | 6245(64.8) | 1578(64.8) | 4668(63.5) |  |
| **Smoking status, n (%)** |  |  |  |  |
| Never smoked | 8237 (84.0) | 2037 (83.5) | 6200 (84.1) | 0.003 |
| Current smoker | 325 (3.3) | 60 (2.5) | 265 (3.6) |  |
| Former smoker | 1244 (12.7) | 341(14.0) | 903 (12.3) |  |
| **Alcohol consumption, n (%)** |  |  |  |  |
| Never | 3937 (40.2) | 959 (39.3) | 2978 (40.5) | 0.708 |
| Monthly or less | 2988 (30.5) | 760 (31.2) | 2228 (30.3) |  |
| 2-3 times per month | 1170 (11.9) | 299 (12.3) | 871 (11.8) |  |
| Once a week or more | 1706 (17.4) | 421 (17.3) | 1285 (17.5) |  |
| **History of diabetes, n (%)** |  |  |  |  |
| No | 9704 (98.6) | 2383 (97.3) | 7321 (99.1) | <0.001 |
| Yes | 135 (1.4) | 67 (2.7) | 68 (0.9) |  |
| **Estimated duration of HIV**<br>**infection (years)** |  |  |  |  |
| Median (25th-75<sup>th</sup> percentiles) | 3.4 (0.2, 8.1) | 5.1(1.1, 9.6) | 2.8 (0.1, 7.4) | <0.001 |
| (Min, Max) | (0.0,25.8) | (0.0, 25.4) | (0.0, 25.8) |  |
| **Any ART use, n (%)** |  |  |  |  |
| No ART | 3194 (32.5) | 956 (39.0) | 2238 (30.3) | <0.001 |
| Yes | 6645 (67.5) | 1494 (61.0) | 5151 (69.7) |  |
| **PI among ART users, n (%)** |  |  |  |  |
| No | 6203(93.9) | 1392(93.7) | 4817(93.9) | 0.799 |
| Yes | 405(6.1) | 93(6.3) | 312(6.1) |  |
| **NRTI among ART users, n**<br>**(%)** |  |  |  |  |
| Yes | 6614(100.0) | 1485(100.0) | 5129(100.0) |  |
| **NNRTI among ART users, n**<br>**(%)** |  |  |  |  |
| No | 302(4.6) | 60(4.0) | 242(4.7) | 0.271 |
| Yes | 6312(95.4) | 1425(96.0) | 4887(95.3) |  |
| INSTI among ART users, n (%) |  |  |  |  |
| No | 6610(99.9) | 1483(99.9) | 5127(100.0) | 0.186 |
| Yes | 4(0.1) | 2(0.1) | 2(0.0) |  |
| ART duration (years) |  |  |  |  |
| Median (25th-75<sup>th</sup> percentiles) | 2.9(0.0, 7.6) | 4.7(0.7, 9.2) | 2.3(0.0, 6.9) | <0.001 |
| (Min, Max) | (0.0, 18.5) | (0.0, 17.6) | (0.0, 18.5) |  |
| CD4 cell count |  |  |  |  |
| Median (IQR) | 373 (184, 573) | 409 (23, 598) | 359 (171, 558) | <0.001 |
| (Min, Max) | (0, 1825) | (0, 1825) | (0, 1753) |  |
| Bp lowering medication, n (%) |  |  |  |  |
| No | 9811(99.7) | 2422 (98.9) | 7389 (100.0) |  |
| Yes | 28 (0.3) | 28 (1.1) |  |  |
| Systolic blood pressure (mmHg) |  |  |  |  |
| Median (25th-75<sup>th</sup> percentiles) | 119.0 (107.0, 134.0) | 147.0 (139.0, 160.0) | 113.0 (103.0, 123.0) | <0.001 |
| (Min, Max) | (47.0, 276) | (93.0, 276) | 47.0, 139.0) |  |
| Diastolic blood pressure (mmHg) |  |  |  |  |
| Median (25th-75<sup>th</sup> percentiles) | 77.0(69.0, 86.0) | 94.0(90.0, 101.0) | 73.0(66.0, 80.0) | <0.001 |
| (Min, Max) | (20.0, 190.0) | (38.0, 190.0) | (20.0, 89.0) |  |
| Body mass index (kg/m²) |  |  |  |  |
| Median (25th-75<sup>th</sup> percentiles) | 24.1(21.4, 27.7) | 25.9(22.8, 30.1) | 23.5(21.0, 27.0) | <0.001 |
| (Min, Max) | (10.4, 87.2) | (10.9, 87.2) | (10.4, 73.5) |  |
| Body mass index categories |  |  |  |  |
| (kg/m²), n (%) |  |  |  |  |
| <18 | 431(4.6) | 47(2.0) | 384(5.5) | <0.001 |
| 18-24.9 | 4943(53.2) | 976(41.7) | 3967(57.1) |  |
| 25-29.9 | 2474(26.6) | 720(30.8) | 1754(25.2) |  |
| 30-35 | 1038(11.2) | 398(17.0) | 640(9.2) |  |
| >35 | 401(4.3) | 199(8.5) | 202(2.9) |  |
| Fasting blood sugar (mg/dl) |  |  |  |  |
| Median (25th-75<sup>th</sup> percentiles) | 88.0(79.0, 97.0) | 90.0(80.0, 99.0) | 87.0(78.0, 96.0) | <0.001 |
| (Min, Max) | (41.0, 860.0) | (47.6, 519.0) | (41.0, 860.0) |  |
| Glucose categories, n (%) |  |  |  |  |
| Normal fasting glucose | 3228(32.8) | 784(32.0) | 2444(33.1) | <0.001 |
| Impaired fasting glucose | 666(6.8) | 200(8.2) | 466(6.3) |  |
| Diabetes / provisional diabetes | 264(2.7) | 102(4.2) | 162(2.2) |  |
| No glucose measure | 5681(57.7) | 1364(55.7) | 4317(58.4) |  |

ART: antiretroviral therapy, BP: blood pressure, INSTI: integrase strand transfer inhibitor, NRTI: nucleoside reverse-transcriptase inhibitor, NNRTI: non-nucleoside reverse-transcriptase inhibitor.

### Prevalence of HTN

A quarter of our cohort (n = 2,450, 25.0%) had hypertension HTN at the time of enrollment into IeDEA (figure 1). The sex-specific prevalence of HTN was 26.6% (95% CI: 25.1–28.1) in men and 23.7% (95% CI: 22.7–24.7) in women for crude prevalence (Figure 2) (P < 0.001); and 23.9% (95% CI: 22.2–25.6) and 20.6% (95% CI: 19.5–21.7) after age-standardization. Overall, 1,328 (13.5%) PLWH had high normal BP, 966 (9.8%) had mild HTN, and 511 (5.2%) grade 2 and 373 (3.8%) grade 3 (Figure 2).

**Figure 1:**
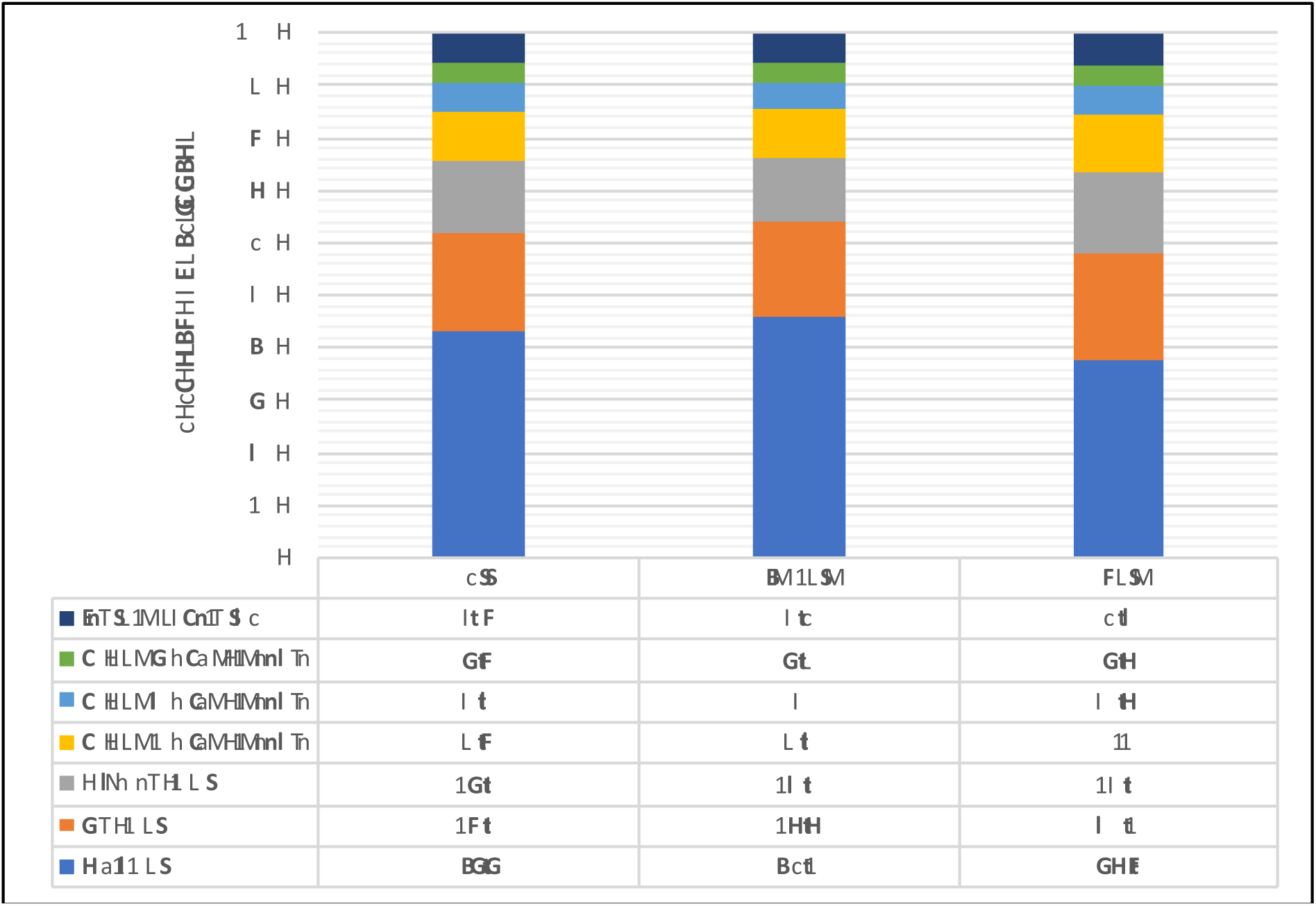
flow chart of the study participants. BP: blood pressure, HTN: hypertension

**Figure 2.**
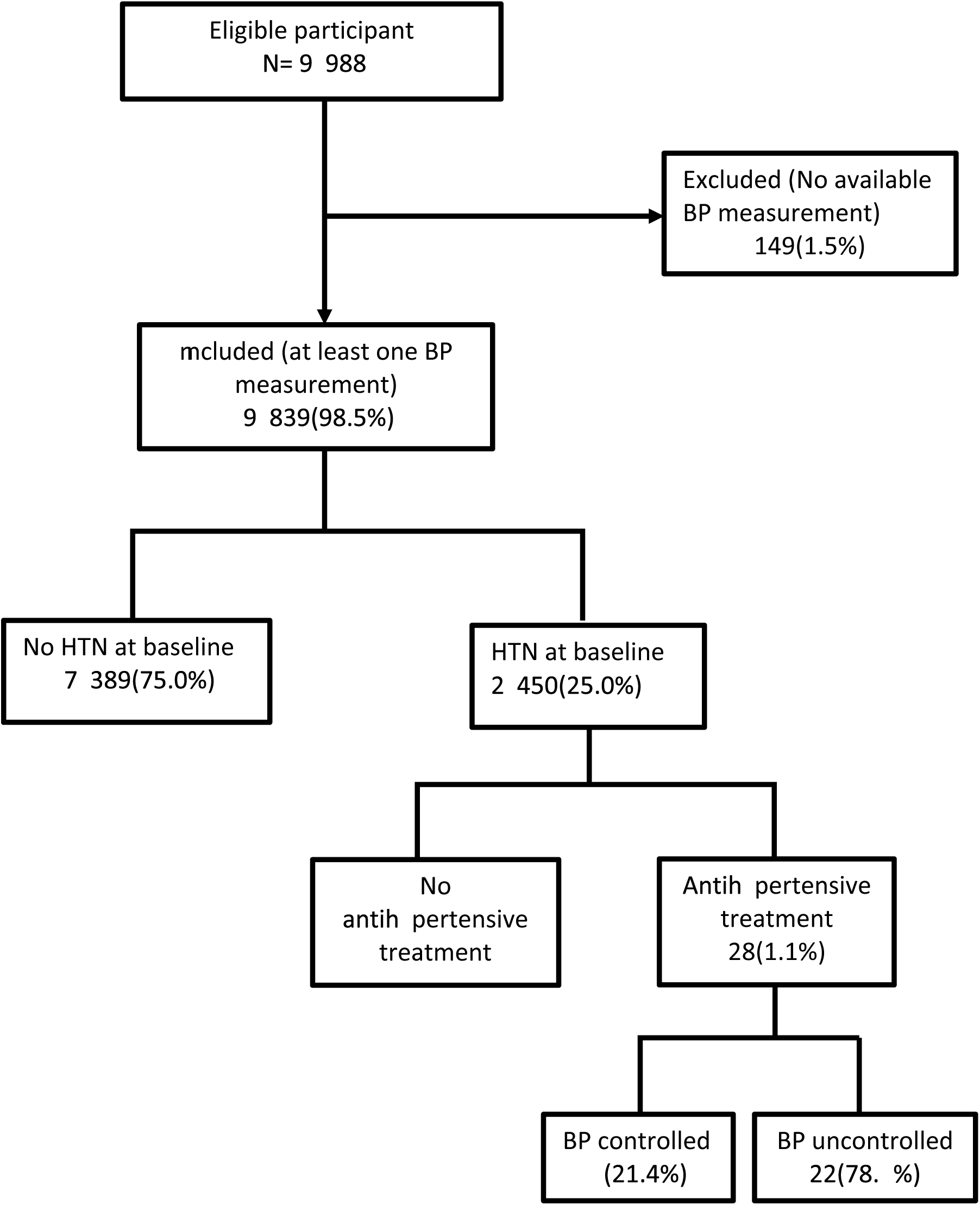
Grade of Blood pressure by 2018 ESC/ESH Guidelines for the overall group (left), female (middle) and male (right).

Table 1 compares a comparison of demographic and clinical characteristics between hypertensive and non-hypertensive PLWH. HTN prevalence increased with increasing age, starting at 9.2% among the < 18 year old rising to 40.5% among those aged > 50 years (p < 0.001) (Table 1). Median diagnosed duration of HIV infection [5.1(1.1, 9.6) vs 2.8(0.1, 7.4) years, p< 0.001] and duration on ART [4.7(0.0, 7.6) vs 2.3(0.0, 6.9), p< 0.001] was longer in participants with HTN than those without HTN. Hypertension was lower among those who did than did not initiate ART (61.0 Vs. 69.7%, p < 0.001). However, among those who initiated ART duration on ART was longer for those with than without HTN [4.7(0.0, 7.6) vs 2.3(0.0, 6.9), p< 0.001]. Based on Loess plots and analyses using indicator variables among ART users once duration reached 2 years, prevalent HTN did not increase further with duration on ART after adjusting for age; meanwhile participants with HTN exhibited higher CD4 count than those without HTN [409 cells/mm^3^ (231 598) vs 359 (171, 558), p< 0.001], Table 1.

### Predictors of hypertension

Table 2 presents OR and 95%CI from crude and multivariable logistic regression models. In multivariable models including age, sex, marital status, BMI, blood glucose level, ART use, ART duration up to 2 years and age, factors significantly associated with HTN were age, sex, BMI, ART use and duration. A 5-year increase in age was independently associated with 28% higher odds of prevalent HTN (adjusted odds ratio (aOR) = 1.28, 95%CI: 1.25–1.32). The adjusted OR of prevalent HTN comparing participants aged > 50 years versus those aged 18–29 years was 4.80 (95%CI: 3.78–6.10). Compared to participants with BMI < 18 kg/m^2^, the adjusted odds of prevalent HTN was 1.87 (95%CI: 1.36–2.57), 2.97 (95%CI: 2.15–4.10), 4.37 (95%CI: 3.11–6.13), and 7.32 (95%CI: 5.04-10.64) respectively among in those with BMI between 18 to 24.9, 25 to 29.9, 30 to 35, and > 35 kg/m^2^. Among those with this measure, higher blood glucose was also independently associated with prevalent HTN with an aOR = 1.47 (95% CI:1.10–1.95) for those with diabetes compared to those with normal blood glucose.

**Table 2.** Predictors of prevalent hypertension.

| **Variable** | **Unadjusted OR\*\***<br>**(95% CI)** | **Adjusted OR**<br>**(95% CI)** |
| --- | --- | --- |
| **Socio-demographic factors** |  |  |
| **Age Group** |  |  |
| 18-29 | Ref | Ref |
| 30-39 | 1.94 (1.56, 2.42) | 1.60 (1.27, 2.02) |
| 40-49 | 3.57 (2.88, 4.43) | 2.65 (2.10, 3.34) |
| 50+ | 6.66 (5.37, 8.26) | 4.80 (3.78, 6.10) |
| **Gender** |  |  |
| Women | Ref | Ref |
| Men | 1.17 (1.06, 1.29) | 1.38 (1.23, 1.54) |
| **Marital status** |  |  |
| Single | Ref | Ref |
| Married | 1.64 (1.46, 1.84) | 1.01 (0.89, 1.16) |
| Live with a partner | 0.87 (0.71, 1.06) | 0.87 (0.71, 1.07) |
| Separated/Divorced | 1.63 (1.35, 1.97) | 0.98 (0.79, 1.20) |
| Widowed | 2.09 (1.83, 2.39) | 1.12 (0.96, 1.32) |
| **Education** |  |  |
| Never went to school | Ref |  |
| Primary | 0.84 (0.72, 0.98) |  |
| Secondary | 0.75 (0.63, 0.88) |  |
| High School or more | 0.91 (0.76, 1.09) |  |
| **Employment** |  |  |
| Unemployed | Ref |  |
| Employed | 1.06 (0.96, 1.17) |  |
| **Cardiovascular risk factors** |  |  |
| **Smoking status** |  |  |
| Never smoked | Ref |  |
| Current smoker | 0.69 (0.52, 0.92) |  |
| Former smoker | 1.15 (1.00, 1.31) |  |
| **Alcohol consumption** |  |  |
| Never | Ref |  |
| Monthly or less | 1.06 (0.95, 1.18) |  |
| 2-3 times per month | 1.07 (0.92, 1.24) |  |
| Once a week or more | 1.02 (0.89, 1.16) |  |
| **Diabetes** |  |  |
| No | Ref |  |
| Yes | 3.03 (2.15, 4.26) |  |
| **Systolic BP (mmHg)** | 6.85 (6.26, 7.49) |  |
| **Diastolic BP (mmHg)** | 14.22 (12.61, 16.03) |  |

| Variable | Unadjusted OR**<br>(95% CI) | Adjusted OR<br>(95% CI) |
| --- | --- | --- |
| **BMI (kg/m<sup>2</sup>)** | 1.09 (1.08, 1.10) |  |
| **BMI (kg/m<sup>2</sup>) categories** |  |  |
| < 18 | Ref | Ref |
| 18 to 24.9 | 2.01 (1.47, 2.74) | 1.87 (1.36, 2.57) |
| 25 to 29.9 | 3.35 (2.45, 4.59) | 2.97 (2.15, 4.10) |
| 30 to 35 | 5.08 (3.66, 7.05) | 4.37 (3.11, 6.13) |
| > 35 | 8.05 (5.61, 11.54) | 7.32 (5.04, 10.64) |
| **Fasting blood sugar (mg/dl)** | 1.02 (1.00, 1.04) |  |
| **Glucose categories** |  |  |
| Normal fasting glucose | Ref | Ref |
| Impaired fasting glucose | 1.34 (1.11, 1.61) | 1.23 (1.00, 1.50) |
| Diabetes / provisional diabetes | 1.96 (1.51, 2.55) | 1.47 (1.10, 1.95) |
| No glucose measure | 0.98 (0.89, 1.09) | 1.09 (0.97, 1.23) |
| **HIV-related factors** |  |  |
| Estimated duration of HIV infection (per year of use) | 1.07 (1.06, 1.08) | 1.15 (1.07-1.24) |
| **Any ART use** |  |  |
| No | Ref | Ref |
| Yes | 0.68 (0.62, 0.75) | 0.70 (0.62, 0.80) |
| **PI among ART users** |  |  |
| No | Ref |  |
| Yes | 1.03 (0.81, 1.31) |  |
| **NNRTI among ART users** |  |  |
| No | Ref |  |
| Yes | 1.18 (0.88, 1.57) |  |
| **INSTI among ART users** |  |  |
| No | Ref |  |
| Yes | 3.46 (0.49, 24.56) |  |
| ART Duration up to 2 years (capped at 2 years, non-ART use considered as 0 years) | 1.10 (1.05, 1.16) | 1.15 (1.07, 1.24) |
| CD4 count | 1.06 (1.03, 1.09) |  |
| Log10-transformed Viral Load | 0.87 (0.77, 0.99) |  |

ART: antiretroviral therapy, BMI: Body Mass Index, BP: blood pressure, NRTI: nucleoside reverse-transcriptase inhibitor, NNRTI: non-nucleoside reverse-transcriptase inhibitor.

Participants on ART at enrollment independently had 30% lower odds of prevalent HTN compared to those not on ART (aOR: 0.70; 95% CI: 0.62–0.80), but there was no significant association between ART regimen (NNRTI or PI) and prevalent HTN (Table 2). Among those using ART, being on ART for up to 2 years was independently associated with higher prevalent HTN (aOR = 1.15/year of ART use, 95% CI:1.07–1.24). Further duration on ART beyond 2 years did not increase the risk of hypertension beyond the fact that those using ART longer tended to be older based on Loess plots (Figure 3). There was a very slow rise in HTN prevalence after 2 years (Figure 3).

**Figure 3.**
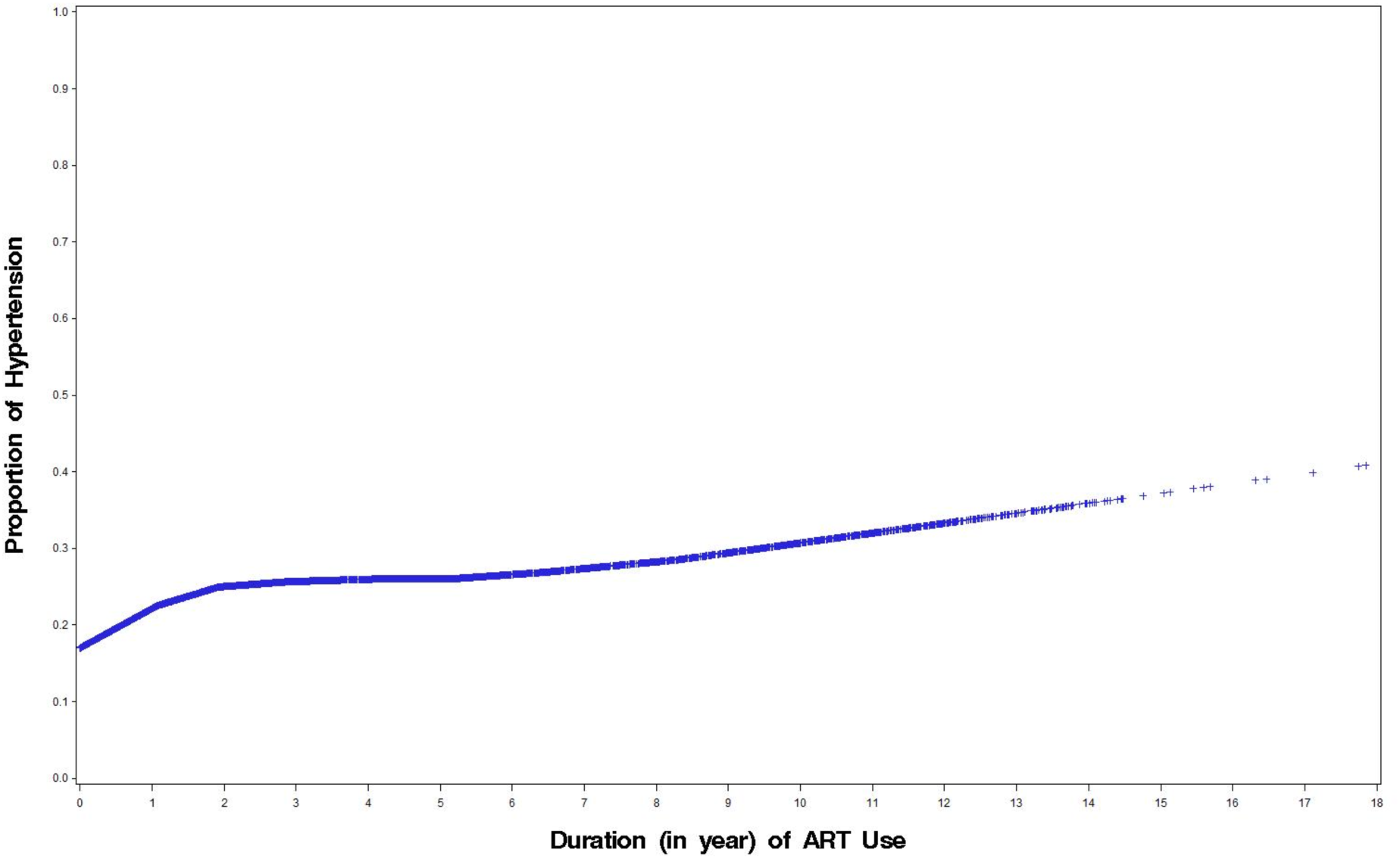
Loess for prevalent hypertension and duration of Antiretroviral therapy (ART) use.

### Treatment and control of HTN

Among participants with HTN, 2422(98.9%) were not receiving any antihypertensive medication; and of the 28 (1.1%) receiving treatment, 6 (21.4%) had a controlled BP (figure 1).

### Discussion

Our study represents the first multicenter and the largest study of HTN in PLWH in west and central Africa and one of the largest in SSA. We found a high crude prevalence of HTN (25%) in PLWH receiving routine care at three HIV clinics across three major cities in the Cameroon, coupled with unacceptably low treatment and control rates. This high prevalence was largely driven by conventional risk factors such as age, sex, and excess weight, and to some extent associated with HIV-related factors such as ART use and duration on ART.

Our findings extend to the population of PLWH in Cameroon and SSA at large, previous observations on the high burden of HTN in the general population across Africa. For instance, we have previously demonstrated in several studies [24–26] that despite being the main cause of stroke, heart and renal disease in Africa, HTN while largely prevalent is still poorly diagnosed, poorly treated and controlled across the continent. Our study clearly highlights the missed opportunity of optimally co-addressing comorbid HTN in PLWH who are already in regular contacts with the health system and at risk of HTN-mediated organ damage.

### Prevalence of hypertension

Over the last decade, three systematic reviews have exclusively focused on the burden of HTN in PLWH. In 2015, Nguyen et al reported prevalence rates ranging from 8% to 45.9% across 13 studies in Africa, of which none were from Cameroon [14]. In a systematic review and metanalysis of 49 studies published during 2011–2016 (10 from Africa including two from Cameroon) with 63,554 participants, Xu et al. reported a global HTN prevalence of 25.2% (95% CI: 21.2% –29.6%) in PLWH [10]. The third and most recent review including 194 studies (396,776 PLWH from 61 countries) reported a 23.6% (95% CI: 21.6–25.5) global prevalence of HTN with little regional variation, 28.1% (95% CI: 24.5–31.9) in Western and Central Europe and North America, with 23.5% (16.6–31.0) in West and Central Africa[27].

Prevalence ranged from 19% to 44.4% across 3 previous Cameroon studies[13][28][29]. It should be noted that these studies all had small samples size ranging from 108 [28] to 311 [29]PLWH and were restricted to single HIV clinics. These shortcomings of Cameroon studies also occurred in studies from other SSA countries and to a certain extent from other regions around the world. For instance, the pooled sample size of PLWH of 17 studies conducted in PLWH in West and Central Africa between 2011–2017 was around 9,010 non randomly selected participants[27].

Apart from the high prevalence, other similarities of HTN burden between our sample of PLWH and the general population lie in the very poor detection, treatment and control rates, as previously described by Mutemwa et al [30] and confirmed by our findings. In 2018, 59.2% of the 8.9 (95% CI: 8.3–9.6) million PLWH with HTN globally were living in Sub-Saharan Africa [27]. In the absence of an integrated HTN and HIV care and no universal health coverage in many SSA countries including Cameroon, PLWH receive ART for free but do not get any BP check and when eventually checked, they do not seek treatment for HTN because of the relatively high cost and poor availability of the medications[31]. This exposes the growing PLWH population to HTN-mediated organ damage and raises strong concerns about the integration of HTN management in HIV clinics including identification of the main drivers of HTN that might be controlled in PLWH.

### Predictors of hypertension

Our findings are generally consistent with previous reports emphasizing the key role of conventional factors in predicting HTN. We found that male sex, higher age and higher BMI increased the odds of prevalent HTN. This is similar to what has been reported in several previous publications both in the general population [26] and in PLWH [32]. With regard to HIV related factors, being on ART at enrolment was associated with 30% lower odds of having HTN, but estimated ART duration up to 2 years was associated with an increased prevalent HTN for both sexes. But this study did not show any significant association between ART regimen (NNRTI, NRTI or PI) and prevalent HTN. The role of HIV-specific risk factors contributing to the development of HTN in PLWH is poorly understood and potentially includes ART exposure and related changes in body composition, immunodeficiency, immune activation and inflammation, as well as effects from antiretroviral therapy itself. Some studies, mostly cross-sectional in design, described higher prevalence rates of HTN in PLWH on ART compared to those not on treatment [29] while others did not find any significant association between ART use and HTN [33][30]. In their metaanalysis and opposite of what we observed, Xu et al[10] described that HTN was 2.7 times more prevalent among ART-exposed PLWH compared with ART naïve. But falling somewhat between, the Strategic Timing of Antiretroviral Treatment clinical trial recently demonstrated that over a follow-up period of 48 months, there were no significant differences in incident HTN between PLWH in the immediate ART group versus deferred treatment group, nor when comparing treated versus untreated HIV infection[34].

We postulate that the protective association of any ART use in our study (aOR = 0.70) may be due to the fact that people initiating ART had more advanced HIV disease i.e. prior to the “treat all” era, had an indication for ART. As proven elsewhere, more advanced HIV disease (WHO stage III/IV, low CD4, and low Hb) is protective against prevalent HTN while low BMI (< 18 kg/m2) is protective against incident hypertension [32]. Indeed, before starting ART, PLWH in SSA usually suffer poor nutrition with severe weight loss, which is associated with a reduction in the BP and may be protective against or otherwise mask diagnosis of HTN [31]. Excess weight stands as one of the main risk factors for development of HTN and weight reduction is key in HTN prevention and control strategies [35]. After starting ART, the initial weight gain and immune reconstitution leading to improved overall health status are likely to explain the increased HTN risk among PLWH, especially after sustained exposure to ART exposure.

Our findings have important implications for policy, practice and research. In the face of this double burden of HIV infection and HTN, integrating HTN’s detection, treatment and control into successful vertical programs for diseases such as HIV/AIDS and tuberculosis as recommended by the Pan African Society of Cardiology[15] is urgently needed to curb the burden. Secondly, healthcare providers should pay more attention to NCD and particularly HTN to prevent the occurrence of cardiovascular events in PLWH. And last but not least, longitudinal studies are needed to understand the pathophysiology of HTN in HIV infection as well as interventional studies to direct innovative and efficient strategies to support prevention and treatment of HTN and HTN-mediated organ damage in PLWH.

### Potential limitations and strengths of our study

Our results should be interpreted considering some limitations. First, there were high levels of missing data for some parameters that could impact on HTN status like WHO stage, creatinine, and hemoglobin. As proven elsewhere, more advanced disease (WHO stage III/IV, low CD4, and low Hemoglobin) is protective against prevalent HTN [32]. Second, we did not have access to HIV-negative populations for comparison. Adding this to the cross-sectional nature of the study, we are unable to draw any causal inference between the diagnosis of HTN and HIV. Considering our recruitment at three large HIV clinics, and targeting nearly 2% of the entire HIV population in the country, these are major strengths and vehicles for accuracy and generalizability. Indeed, in several previous published studies of the burden of HTN in Cameroon, we and others have demonstrated that HTN occurs around the fifth decade in the general population and affects 30% of Cameroonians older than 21 years of age [24,26,37], roughly only 5% higher than the estimated 25% we reported in younger population of PLWH and consistent with reports from other robust studies [32] and a contemporary systematic review metanalysis [27]. Finally, we may have underestimated the proportion of participants with HTN receiving treatment, if that treatment was occurring in another location.

### Conclusions

In conclusion, our study found that about one of every four PLWH in Cameroon treated at three major clinics has HTN, which is largely under-recognized and under-treated, exposing this specific population group to HTN-mediated organ damages. Drivers of HTN in PLWH include both conventional risk factors like older age, male sex and excess weight as well as specific HIV-related factors including ART exposure and estimated duration of ART exposure. These findings should motivate the integration of HTN services at HIV clinics to stop the growing burden of NCDs among PLWH.

## Data Availability

Data for the Central Africa International Epidemiology to Evaluate AIDS (CA-IeDEA) is available upon request.

## Author contributions

AD, AA, MY, DN, and KA conceived the study and acquired the funding.

AD, RA, WPY, DNN, KTN, VPE, ZKBAC operationalized and supervised data collection

AD, DH, HYK, QS, KA, MY, DN and APK analyzed the data and drafted the manuscript.

All co-authors substantially revised the manuscript and approved the submission.

## Acknowledgments list

Nimbona Pélagie, Association Nationale de Soutien aux Séropositifs et Malade du Sida (ANSS), Burundi; Patrick Gateretse, Jeanine Munezero, Valentin Nitereka, Théodore Niyongabo, Christelle Twizere, Centre National de Référence en Matière de VIH/SIDA, Burundi; Hélène Bukuru, Thierry Nahimana, Centre de Prise en Charge Ambulatoire et Multidisciplinaire des PVVIH/SIDA du Centre Hospitalo-Universitaire de Kamenge (CPAMP-CHUK), Burundi; Elysée Baransaka, Patrice Barasukana, Eugene Kabanda, Martin Manirakiza, François Ndikumwenayo, CHUK/Burundi National University, Burundi; Jérémie Biziragusenyuka, Ange Marie Michelline Munezero, Centre de Prise en Charge Ambulatoire et Multidisciplinaire des PVVIH/SIDA de l’Hôpital Prince Régent Charles (CPAMP-HPRC), Burundi; Tabeyang Mbuh, Kinge Thompson Njie, Edmond Tchassem, Kien-Atsu Tsi, Bamenda Hospital, Cameroon; Rogers Ajeh, Mark Benwi, Marc Lionel Ngamani, Victorine Nkome, Grace Toutou, Clinical Research Education and Consultancy (CRENC), Cameroon; Anastase Dzudie, CRENC and Douala General Hospital, Cameroon; Akindeh Mbuh, CRENC and University of Yaoundé, Cameroon; Djenabou Amadou, Amadou Dodo Balkissou, Eric Ngassam, Eric Walter Pefura Yone, Jamot Hospital, Cameroon; Alice Ndelle Ewanoge, Norbert Fuhngwa, Ernestine Kendowo, Chris Moki, Denis Nsame Nforniwe, Limbe Regional Hospital, Cameroon; Catherine Akele, Akili Clever, Faustin Kitetele, Patricia Lelo, Martine Tabala, Kalembelembe Pediatric Hospital, Democratic Republic of Congo; Cherubin Ekembe, Didine Kaba, Kinshasa School of Public Health, Democratic Republic of Congo; Merlin Diafouka, Martin Herbas Ekat, Dominique Mahambou Nsonde, CTA Brazzaville, Republic of Congo; Adolphe Mafoua, Massamba Ndala Christ, CTA Pointe-Noire, Republic of Congo; Jules Igirimbabazi, Nicole Ayinkamiye, Bethsaida Health Center, Rwanda; Providance Uwineza, Emmanuel Ndamijimana, Busanza Health Center, Rwanda; Emmanuel Habarurema, Marie Luise Nyiraneza, Gahanga Health Center, Rwanda; Marie Louise Nyiransabimana, Liliane Tuyisenge, Gikondo Health Center, Rwanda; Christian Shyaka, Catherine Kankindi, Kabuga Health Center, Rwanda; Bonheur Uwakijijwe, Marie Grace Ingabire, Kicukiro Health Center, Rwanda; Jules Ndumuhire, Marie Goretti Nyirabahutu, Masaka Health Center, Rwanda; Fred Muyango, Jean Christophe Bihibindi, Nyagasambu Health Center, Rwanda; Oliver Uwamahoro, Yvette Ndoli, Nyarugunga Health Center, Rwanda; Sabin Nsanzimana, Placidie Mugwaneza, Eric Remera, Esperance Umumararungu, Gallican Nshogoza Rwibasira, Dominique Savio Habimana, Rwanda Biomedical Center, Rwanda; Josephine Gasana, Faustin Kanyabwisha, Gallican Kubwimana, Benjamin Muhoza, Athanase Munyaneza, Gad Murenzi, Francoise Musabyimana, Francine Umwiza, Charles Ingabire, Patrick Tuyisenge, Alex M. Butera, Jules Kabahizi, Ephrem Rurangwa, Rwanda Military Hospital, Rwanda; Rosine Feza, Eugenie Mukashyaka, Shyorongi Health Center, Rwanda; Chantal Benekigeri, Jacqueline Musaninyange, WE-ACTx Health Center, Rwanda.

## Coordinating and Data Centers

Adebola Adedimeji, Kathryn Anastos, Madeline Dilorenzo, Lynn Murchison, Jonathan Ross, Marcel Yotebieng, Albert Einstein College of Medicine, USA; Diane Addison, Ellen Brazier, Heidi Jones, Elizabeth Kelvin, Sarah Kulkarni, Denis Nash, Matthew Romo, Olga Tymejczyk, Institute for Implementation Science in Population Health, Graduate School of Public Health and Health Policy, City University of New York (CUNY), USA; Batya Elul, Columbia University, USA; Xiatao Cai, Allan Dong, Don Hoover, Hae-Young Kim, Chunshan Li, Qiuhu Shi, Data Solutions, USA; Kathryn Lancaster, The Ohio State University, USA; Mark Kuniholm, University at Albany, State University of New York, USA; Andrew Edmonds, Angela Parcesepe, Jess Edwards, University of North Carolina at Chapel Hill, USA; Olivia Keiser, University of Geneva; Stephany Duda; Vanderbilt University School of Medicine, USA; April Kimmel, Virginia Commonwealth University School of Medicine, USA.

## Supporting institutions

Professor Zoung-Kany Bisseck Anne Cecile, Division of operational health research (DROS), Ministry of Public Health (MSP), Cameroon, Dr Elat Mfetam Jean Bosco and Dr Bonono Leonard, National AIDS Control Committee (NACC), MSP, Cameroon Ministry of Public Health, Albert Einstein College of Medicine, USA.

## Participants to the study

All PLWH for attending the clinics for their continued trust in our health care system

## Funding acknowledgement

Research reported in this publication was supported by the National Institutes of Health’s National Institute of Allergy and Infectious Diseases (NIAID), the *Eunice Kennedy Shriver* National Institute of Child Health & Human Development (NICHD), the National Cancer Institute (NCI), the National Institute on Drug Abuse (NIDA), the National Heart, Lung, and Blood Institute (NHLBI), the National Institute on Alcohol Abuse and Alcoholism (NIAAA), the National Institute of Diabetes and Digestive and Kidney Diseases (NIDDK), the Fogarty International Center (FIC), the National Library of Medicine (NLM), and the Office of the Director (OD) under Award Number U01AI096299 (Central Africa-IeDEA). The content is solely the responsibility of the authors and does not necessarily represent the official views of the National Institutes of Health.

